# Adolescents and adults with FOXP1 syndrome show high rates of anxiety and externalizing behaviors but not psychiatric decompensation or skill loss

**DOI:** 10.1101/2025.01.04.24318923

**Authors:** Tess Levy, Hailey Silver, Renee Soufer, Audrey Rouhandeh, Alexander Kolevzon, Joseph D. Buxbaum, Paige M. Siper

**Affiliations:** Seaver Autism Center for Research and Treatment, Department of Psychiatry, Icahn School of Medicine at Mount Sinai, New York, NY 10029, USA; Department of Psychiatry, Icahn School of Medicine at Mount Sinai, New York, NY 10029, USA; Ferkauf Graduate School of Psychology. Yeshiva University. New York, NY 10461, USA; The Mindich Child Health and Development Institute, Icahn School of Medicine at Mount Sinai, New York, NY 10029, USA; Department of Pediatrics, Icahn School of Medicine at Mount Sinai, New York, NY 10029, USA; Friedman Brain Institute, Icahn School of Medicine at Mount Sinai, New York, NY, 10029, USA; Department of Genetics and Genomic Sciences, Icahn School of Medicine at Mount Sinai, New York, NY, 10029, USA; Department of Neuroscience, Icahn School of Medicine at Mount Sinai, New York, NY, 10029, USA

**Keywords:** FOXP1, psychiatric features

## Abstract

**Background:** FOXP1 syndrome is a genetic neurodevelopmental disorder associated with complex clinical presentations including global developmental delay, mild to profound intellectual disability, speech and language impairment, autism traits, attention-deficit/hyperactivity disorder (ADHD), and a range of behavioral challenges.. To date, much of the literature focuses on childhood symptoms and little is known about the FOXP1 syndrome phenotype in adolescence or adulthood.

**Methods:** A series of caregiver interviews and standardized questionnaires assessed psychiatric and behavioral features of 20 adolescents and adults with FOXP1 syndrome. Interviews captured change in various psychiatric manifestations over time. Medication, social, educational, and vocational history was collected and visual analog scales measured top caregiver concerns during childhood and adolescence/adulthood.

**Results:** Anxiety and externalizing behaviors were common in this cohort and psychiatric features, such as psychosis or bipolar symptoms, were present in two participants. There was no reported regression or loss of skills, early in development or during adolescence/adulthood. Caregivers reported continued development in adaptive skills even into adolescence/adulthood. Medication use was common particularly for features of ADHD, although multiple trials were required for some participants to achieve benefit. Standardized assessments accurately picked up on externalizing symptoms and were less sensitive to internalizing symptoms. Educational setting varied up until late elementary school and gradually shifted to special education. Cognitive and developmental concerns were reported as primary during childhood and independence/safety and housing concerns became top concerns by adolescence/adulthood. Caregivers reported continued development in adaptive skills even into adulthood.

**Conclusions:** Taken together, results are reassuring, with many families reporting their adolescent and adult children continued to gain skills over time, particularly related to increased independence in communication and personal care. There were no reports of developmental regression, neuropsychiatric decompensation or catatonia.

## BACKGROUND

FOXP1 syndrome is a rare genetic disorder caused by heterozygous sequence variants in — or deletions of — the *FOXP1* gene*. FOXP1* codes for the Forkhead box protein P1 (Foxp1), a member of the Fox family of transcription factors. Along with Foxp2, which has been implicated in human speech and language (1), Foxp1 is involved in transcriptional regulation and is highly expressed in the brain. Though the exact function of Foxp1 within the human brain is not completely resolved, animal models have pointed to hippocampal and striatal functions. In mice with brain-specific *Foxp1* deletion, alterations in the developing striatum and hippocampus were identified, along with behavioral features of cognitive and social deficits (2). Additionally, studies using ubiquitous (whole-body) mouse knockout models and human neural models have demonstrated a role in regulation of genes involved in striatal development (3). Pathogenic alterations of *FOXP1* within FOXP1 syndrome are varied, with protein-truncating variants (nonsense, frameshift, splice site), and missense variants all being reported. Deletion copy number variants including the *FOXP1* gene have been shown to result in a similar phenotype as *FOXP1* sequence variants (4–6), indicating that haploinsufficiency as one mechanism of disease. Investigation into missense variation has suggested there may be dominant negative effects of some mutations, including p.R465G and p.R514C (7), and loss of function of others, including p.E482K (8), although this requires more study in native systems. Recurrent variants exist, such as the p.R525Q missense mutation (9).

The clinical presentation of FOXP1 syndrome has largely been characterized from studies of youth. These studies show that, FOXP1 syndrome is associated with global developmental delay, intellectual disability, autism traits, and attention-deficit/hyperactivity disorder (ADHD) (10–15). Common medical features include gastrointestinal problems, hypotonia, and vision abnormalities, among many other medical comorbidities with varying frequency. Intellectual disability (ID) is present in most cases and generally falls in the mild to moderate range (10, 11). Language impairment is present in all cases and can range from individuals with few to no words to those achieving fluent speech. However, language acquisition is significantly delayed, and complex speech is rarely obtained (e.g., using *and* or *but* to combine clauses; speaking in detail about the past or future). In spite of these insights, little is known about the natural history of the syndrome or trajectories into adolescence and adulthood.

Reports from other genetic syndromes shed light on significant challenges emerging around the adolescent period. For example, Phelan-McDermid syndrome (PMS), which is also characterized by high rates of intellectual disability, autism spectrum disorder (ASD)/autism traits, language impairment, and medical comorbidities, is associated with risk of significant skill regression around and after puberty and the development of psychiatric disorders such as bipolar disorder and catatonia in certain patients (16). These changes in PMS can be quite pronounced and include significant loss of language, motor skills, and self-help skills. A subset of individuals with DLG4-related neurodevelopmental disorder are also reported to develop bipolar disorder, depression, and hallucinations around adolescence (17). In 22q11.2 deletion syndrome (DiGeorge syndrome), up to one third of individuals develop psychotic features in adolescence that resemble schizophrenia (18). Individuals with Down Syndrome and Kleefstra syndrome are also at a higher risk for psychotic features (19, 20).

Complex psychiatric manifestations reduce quality of life for individuals and their caregivers and may be difficult to diagnose and treat in those with ID. Concerning post-pubertal findings from these other neurogenetic syndromes prompted the current study to assess evidence for the emergence of regression, or behavioral, and psychiatric changes in FOXP1 syndrome and to examine developmental trajectory during and after puberty. The information gathered from this study is of high importance to the FOXP1 community and is critical for the design of natural history studies and future clinical trials.

## METHODS

### Participants and procedures

Participants included 20 individuals with FOXP1 syndrome (12 females, 8 males) between the ages of 13-35 (*M=*19.8 *SD=*6.2). Of these participants, five were previously known to our center and previously published (11). All participants had a likely pathogenic or pathogenic variant in *FOXP1*, confirmed by a genetic counselor using ACMG-AMP criteria (21). Eligibility criteria required that participants had begun or completed puberty as confirmed by a standard Tanner Stage Checklist and had a parent able to be the informant for the study. All caregiver interviews were administered remotely via Zoom and questionnaires were completed via REDCap surveys, with the exception of the Child or Adult Behavior Checklist which were administered through the Achenbach System of Empirically Based Assessment directly. Descriptive statistics and figure creation were made in Rstudio. This study was approved by Mount Sinai’s Program for the Protection of Human Subjects. Informed consent was obtained from parents or legal guardians.

### Measures

*Clinical Evaluation.* A clinical interview was conducted with caregivers to obtain history of present illness, birth and developmental history, medical history, psychiatric history, social history, educational history, vocational history, and family history. A medication log was also completed by caregivers to gather current and past medication history, perceived efficacy, and side effects.

*Caregiver Interview for Psychiatric Illness in Persons with ID (CIPIPID).* The CIPIPID is a semi-structured caregiver interview used to assess psychiatric symptoms in people with ID in the following areas: depression, mania, catatonia, disoriented/psychotic behavior, anxiety, self-injury, and aggression (16). The CIPIPID is intended to capture these symptoms in individuals with intellectual and language impairment.

*Diagnostic and Statistical Manual of Mental Disorders Fifth Edition (DSM-5) Cross-Cutting Symptom Measure* was administered to caregivers by a clinician in an interview format to ensure consistent rating of items (i.e., due to limited language ability in some participants)(22). This measure screened for somatic symptoms, inattention, depression, anger and irritability, mania, anxiety, psychosis, repetitive thoughts and behaviors, and suicidal behaviors that had occurred over the past two weeks. For all endorsed items, frequency is measured on a Likert scale.

*Early Skill Attainment and Loss (Early Skills)*. The Early Skill Attainment and Loss interview was administered to caregivers to capture age of skill acquisition in the following domains: social, motor, daily living, and language. The presence of regression and any subsequent skill reattainment was obtained. Regression was defined as the loss of a skill after the skill had been present and used reliably for at least three months.

*Vineland Adaptive Behavior Scales, 3rd Edition - Comprehensive Interview Form (Vineland-3)*. Adaptive functioning was measured using Vineland-3 and administered to caregivers (23). Domain scores (Communication, Daily Living, Socialization, Motor, Maladaptive Behavior) are measured in standard scores (mean = 100; SD = 15). Subdomain scores are measured in v-scale scores (mean = 15; and SD = 3) and age equivalents (presented in years).

*Sensory Assessment for Neurodevelopmental Disorders (SAND) - Interview.* The SAND measures hyperreactivity, hyporeactivity, and seeking across visual, tactile, and auditory domains (24). This standardized assessment combines a clinician-administered observation and corresponding caregiver interview. Due to the virtual nature of this study, only the interview was administered.

*Short Sensory Profile 2 (SP-2)*. The Short SP-2 is a caregiver questionnaire that measures sensory processing and behavioral sensory responses based on experiences in everyday settings (25).

*Aberrant Behavior Checklist (ABC).* The Aberrant Behavior Checklist is a caregiver questionnaire that captures problem behaviors in individuals with developmental disorders (26). Raw scores and T-scores are generated from each of the five domains: irritability, social withdrawal/lethargy, hyperactivity, stereotypy, and inappropriate speech. A T-score of 65 or above, or 1.5 standard deviations above the mean, was considered elevated.

*Child Behavior Checklist (CBCL) or Adult Behavior Checklist (ABCL).* The Child and Adult Behavior Checklists are caregiver questionnaires that measure problem behaviors (27). Domains include syndrome scales, internalizing, externalizing, total problems, and DSM-oriented scales. Nine caregivers completed the CBCL for ages 6 to 18 and 10 caregivers completed the ABCL for ages 18 and older.

*Visual Analog Scale (VAS).* Caregivers reported their top three concerns on a VAS from 0 to 100 (severity). Ratings were obtained for (1) how concerned caregivers were in early childhood and (2) how concerned caregivers are now. Caregivers then reported top concerns at their child’s current age and rated severity from 0-100. The VAS is available as Additional File 1.

## RESULTS

### Participants Demographics

Participants were between 13 and 35 years old (*Mage 19*.8 ± 6.2), including 12 females and 8 males. Eighteen participants had a likely pathogenic or pathogenic sequence variant in *FOXP1,* the remaining two participants had a 3p13 deletion including the *FOXP1* gene. There were seven missense variants, seven frameshift variants, two nonsense variants, and two intronic/splice variants. Genetic alterations were confirmed to be *de novo* in 13 cases, one participant’s parent had low level mosaicism of the *FOXP1* variant (Arg514His), and in six cases parental testing was not completed. All participants had begun puberty as confirmed by a Tanner Stage Checklist, with most in or past stage 4 (28, 29). All females had begun menstruation at the time of participation. There were 18 white participants, one of whom was Hispanic/Latinx, one Black participant, and one Asian participant.

### Developmental Milestones

Participants experienced global developmental delays. Sitting was achieved at an average age of 13 months (± 12 months) and walking at 22 months (± 5 months). All participants walked independently and 18 crawled prior to walking. There were no reports of motor skill regression. Regarding language, all 20 participants achieved single words, 19/20 achieved phrase speech, and 11/20 spoke in full sentences. Average age of first single words was 22 months (± 13 months), two-word phrases were 43 months (± 27 months), and fluent speech was 6.9 years (± 4.2 years). A history of language delay/impairment was reported in all but one participant.

Articulation problems were common and intelligibility issues were reported in many participants who spoke in sentences. Regarding language changes around puberty, one parent reported episodes of less intelligible speech lasting for several months at a time, requiring intermittent speech therapy. One parent reported the frequency of language use declined during adolescence, despite improved complexity of speech. Several parents of individuals who spoke in full sentences reported continued improvement in the length and complexity of speech post-puberty including two descriptions of improvement up until their late teens and early 20s. Those with less speech early on (i.e., single words/occasional phrases) reported plateaus in language prior to adolescence.

Regarding continence, 14/20 achieved bladder control and 18/20 achieved bowel control. Bladder control was achieved, on average, at 9 years, 5 months (± 61 months) and bowel control at an average age of 5 years, 1 month (± 20 months). Despite this, many continue to have accidents and/or require assistance or reminders as described in the Adaptive Functioning section below. There was no report of regression in any domain occurring around puberty. Most parents reported continued development over time, particularly growth related to independence in activities of daily living.

### Adaptive Functioning

Results from the Vineland-3 indicate significant deficits in adaptive functioning (Table 1). Average standard scores for the Communication, Daily Living Skills, and Socialization domains all fell below the 1^st^ percentile.

**Table 1.** Vineland-3 Interview Scores. Standard scores are provided for domain scores; V-scale scores, age equivalents, and growth scale values are provided for subdomain scores. Internalizing and Externalizing behavior only have V-scale scores and do not have age equivalents or growth scale values. Values are presented as mean (standard deviation).

|  | Standard Score | V-Scale Score | Age Equivalent (years) | Growth Scale Value |
| --- | --- | --- | --- | --- |
| Adaptive Behavior Composite | 36.95 (14.00) |  |  |  |
| Communication | 31.75 (12.56) |  |  |  |
| Receptive |  | 5.3 (3.79) | 3.79 (2.68) | 118.65 (16.86) |
| Expressive |  | 2.65 (2.58) | 3.37 (1.13) | 151.8 (16.45) |
| Written |  | 3.4 (2.84) | 5.93 (1.20) | 88.9 (21.06) |
| Daily Living Skills | 33.50 (18.37) |  |  |  |
| Personal |  | 3.6 (3.39) | 4.80 (2.85) | 123.3 (17) |
| Domestic |  | 4.65 (3.92) | 5.75 (3.55) | 47.95 (19.2) |
| Community |  | 4.65 (2.78) | 6.80 (3.13) | 62.05 (17.97) |
| Socialization | 43.25 (17.63) |  |  |  |
| Interpersonal |  | 5.85 (3.1) | 2.89 (1.43) | 98.40 (18.35) |
| Play and Leisure |  | 6.25 (3.48) | 4.88 (3.18) | 101.3 (25.51) |
| Coping |  | 5.15 (3.38) | 3.73 (2.24) | 60.05 (19.66) |
| Internalizing Behavior |  | 19.44 (2.01) |  |  |
| Externalizing Behavior |  | 20.11 (2.56) |  |  |

In the Daily Living Skills domain, 13 of 20 consistently dress themselves and another three do so sometimes. Eleven shower independently (10 usually, 1 sometimes) and 10 wash their hair independently (7 usually, 3 sometimes). Eight participants were reported to brush their teeth independently. Ten participants were reported to show safety awareness around hot and sharp objects (9 usually, 1 sometimes). In the kitchen, seven use kitchen utensils and appliances to prepare food (6 usually, 1 sometimes), and 11 were reported to do simple household chores (9 usually, 2 sometimes). Many rely on assistance, prompting, or reminders for other activities of daily living.

In terms of toileting, eight caregivers reported independence in toileting during both night and day, with two reporting that they are sometimes independent in both. Of those who were toilet trained and out of diapers/pull ups, 61% continued to have accidents. The following information was provided during the clinical interview: medicated for accidents (n=1), several daytime accidents a month (n=1), regular nighttime and occasional daytime accidents (n=1), bowel accidents when nervous (n=1), bowel accidents when laughing hard (occurs every ∼3 weeks) (n=1), occasional accidents (n=4), requires pull up at night (n=1), still defecates in a diaper and is cleaned by an adult (n=1). Many participants were reported to require help or reminders to wipe. Only one individual was fully toilet trained without accidents and the skill was attained by age three.

### Education and Vocation

Classroom setting was reported by caregivers for each stage of education (Figure 1). There was a gradual shift towards special education and away from mainstream and inclusion classrooms. No participants remained in a mainstream class after elementary school without full time support.

**Figure 1.**
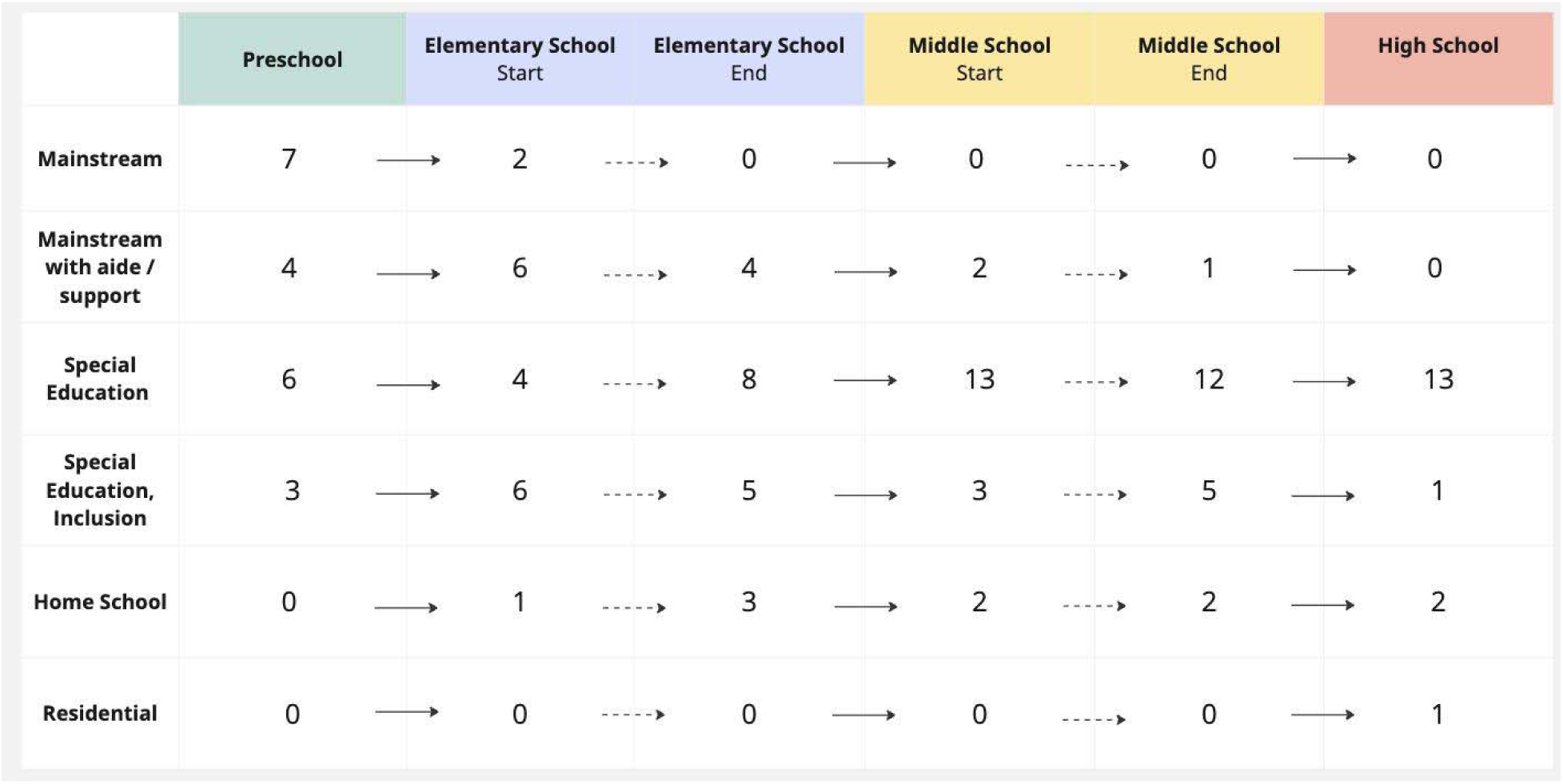
Educational setting of participants. Education setting in the cohort. participant. Mainstream with aide / support included individuals with a 1:1 aide, modified curriculum, and pull-out supports. Three individuals were not yet in high school.

For those who began elementary school in a mainstream class, grades 3-4 were most commonly reported as times of transition where increased support was necessary.

Eighteen of 20 participants received speech therapy at one point in time; 15/20 during early intervention, 16/20 in pre-kindergarten/kindergarten, 18/20 in elementary school, 14/20 in middle school, and 12/17 in high school (3 participants were not yet in high school). Physical therapy was received by 17/20 participants; 5/20 in early intervention, 5/20 in pre-kindergarten/kindergarten, 11/20 in elementary school, 9/20 in middle school, and 4/17 in high school. Nineteen of 20 participants received occupational therapy at one point in time; 17/20 in early intervention, 18/20 in pre-kindergarten/kindergarten, 17/20 in elementary school, 12/20 in middle school, and 7/17 in high school. Six of 20 participants received ABA therapy, 2 in early intervention, 2 in pre-kindergarten/kindergarten, 2 in elementary school, 4 in middle school, and 2 in high school.

In terms of vocation, five participants had paid jobs; the youngest participant with a paid job was in their teens. Hours per week ranged from 5 to 27 and job types included yard and garden work, grocery or home improvement stores, clerical work, and maintenance/cleaning. Four of five participants had independent responsibilities at their jobs. Caregivers of three participants reported that their children expressed enjoyment of their work and two reported they assumed their children enjoyed the work due to absence of complaints. Common reasons for those without a job included not having the independence to hold a job or requiring constant 1:1 support (n=7), being too busy with school or too young (n=4), and unwillingness to complete non-preferred activities (n=2). Seven participants were preparing to have a job, and 11 caregivers reported a desire for their child to hold a job one day.

### Medical History

Medical features reported in this cohort are located in Table 2. Strabismus (n=5) and esotropia (n=2) were reported ocular findings and six participants required corrective lenses. Sleep problems were commonly reported. On the DSM-5 checklist, which asks specifically about symptoms experienced over the past two weeks, 7/20 reported severe sleep problems (nearly every day), 2/20 reported several days per week, 3/20 reported less than a day or two per week, and 8/20 reported none. Parent descriptions regarding current and past sleep problems collected during the clinical evaluation indicated three groups: chronic sleep problems (n=5), sleep problems that improved with age (n=9; note several improved due to ongoing medication), and rare to none (n=5). In all those with chronic sleep problems, and most with improved sleep problems, caregivers reported difficulty with both sleep onset and maintenance. Reported congenital anomalies included cardiac (2/4), spina bifida (1/4), and one participant had hip dysplasia (1/4), and contractures. Gastrointestinal problems included reflux, constipation, and diarrhea. Recurrent infections included ear infections, urinary tract infections, and skin infections in one participant each.

**Table 2.** Medical Features.

| Features | Proportion |
| --- | --- |
| Vision/ocular problems | 13/20 |
| Sleep problems | 12/20 |
| Abnormal head circumference (micro/macrocephaly) | 7/20 |
| Hypotonia | 6/20 |
| Congenital anomalies | 4/20 |
| Seasonal allergies | 4/20 |
| Gastrointestinal problems | 3/20 |
| Recurrent infections | 3/20 |
| Hearing problems | 2/20 |
| Seizure disorder | 2/20 |
| Limited endurance | 2/20 |

### Psychiatric and Behavioral Presentations

#### Internalizing Symptoms

During the psychiatric evaluation, anxiety symptoms were reported in 17/20 participants and 12 were diagnosed with an anxiety disorder by a medical professional (Figure 2, Table 3). One parent reported their child’s anxiety resolved with age, seven reported decreased anxiety over time, and three reported worsening anxiety with age. The remaining six parents reported relatively stable symptoms and they were generally reported as mild (e.g., anxiety only when out of routine or related to unexpected circumstances). Somatic symptoms were reported in one individual who was also reported to have an anxiety disorder; this individual had higher adaptive functioning among the cohort (i.e., maintained a longstanding paid job). There was one report of an individual with anxiety, depression and suicidal ideation along with worsening social skills, aggression, and irritability over time. This individual also experienced sleep problems and attention problems. There was no report of substance abuse, and most participants did not partake in any substance use.

**Figure 2.**
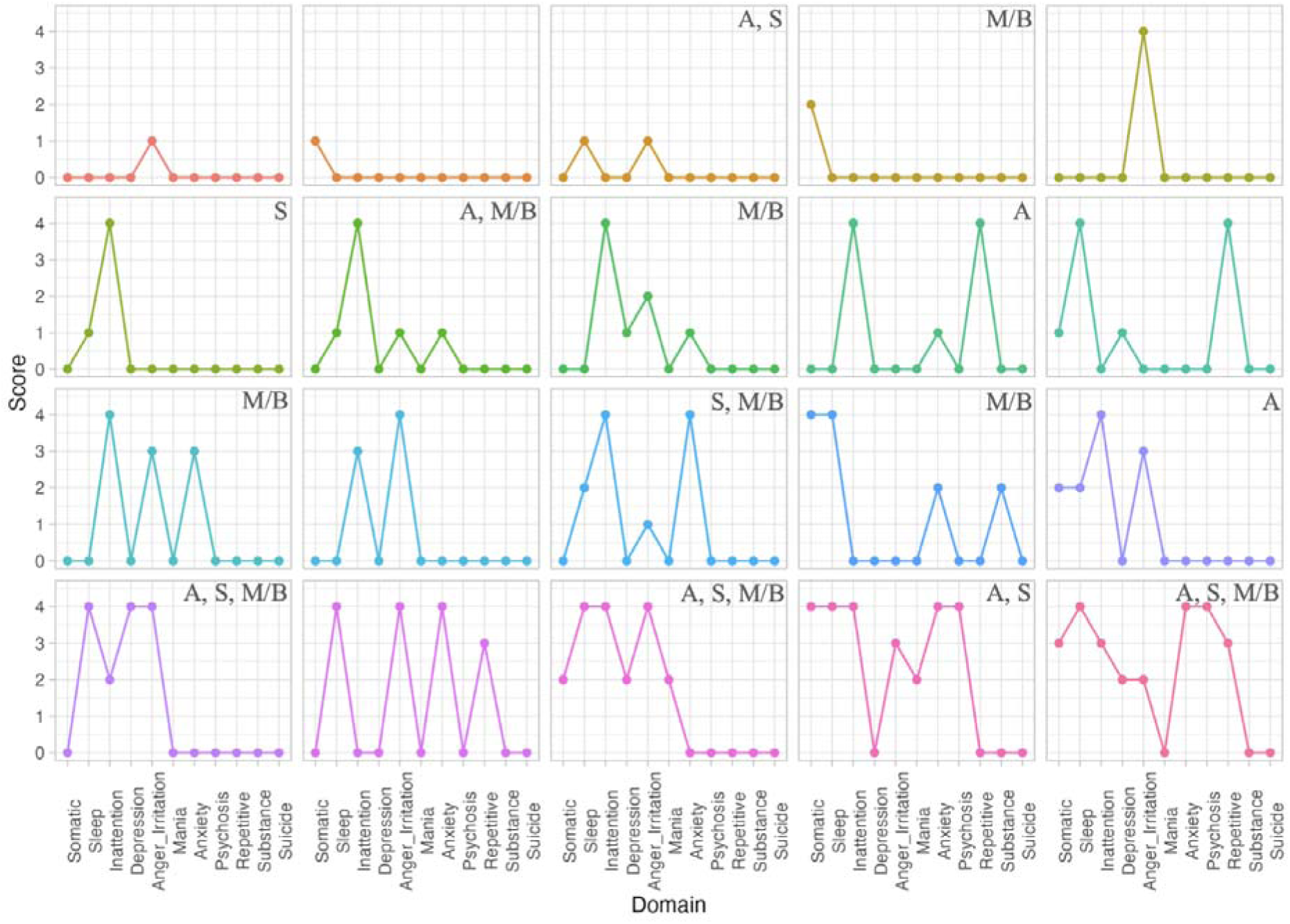
DSM-5 Cross Cutting Interview Results. Top score within each DSM-5 domain as part of the cross-cutting interview. Participants are ordered from least to most domains with scores two or higher. Letters in the top right corner indicate current treatment with psychiatric medications: A: medication for ADHD, S: medication for sleep, M/B: medication for mood / behavior.

**Table 3.**
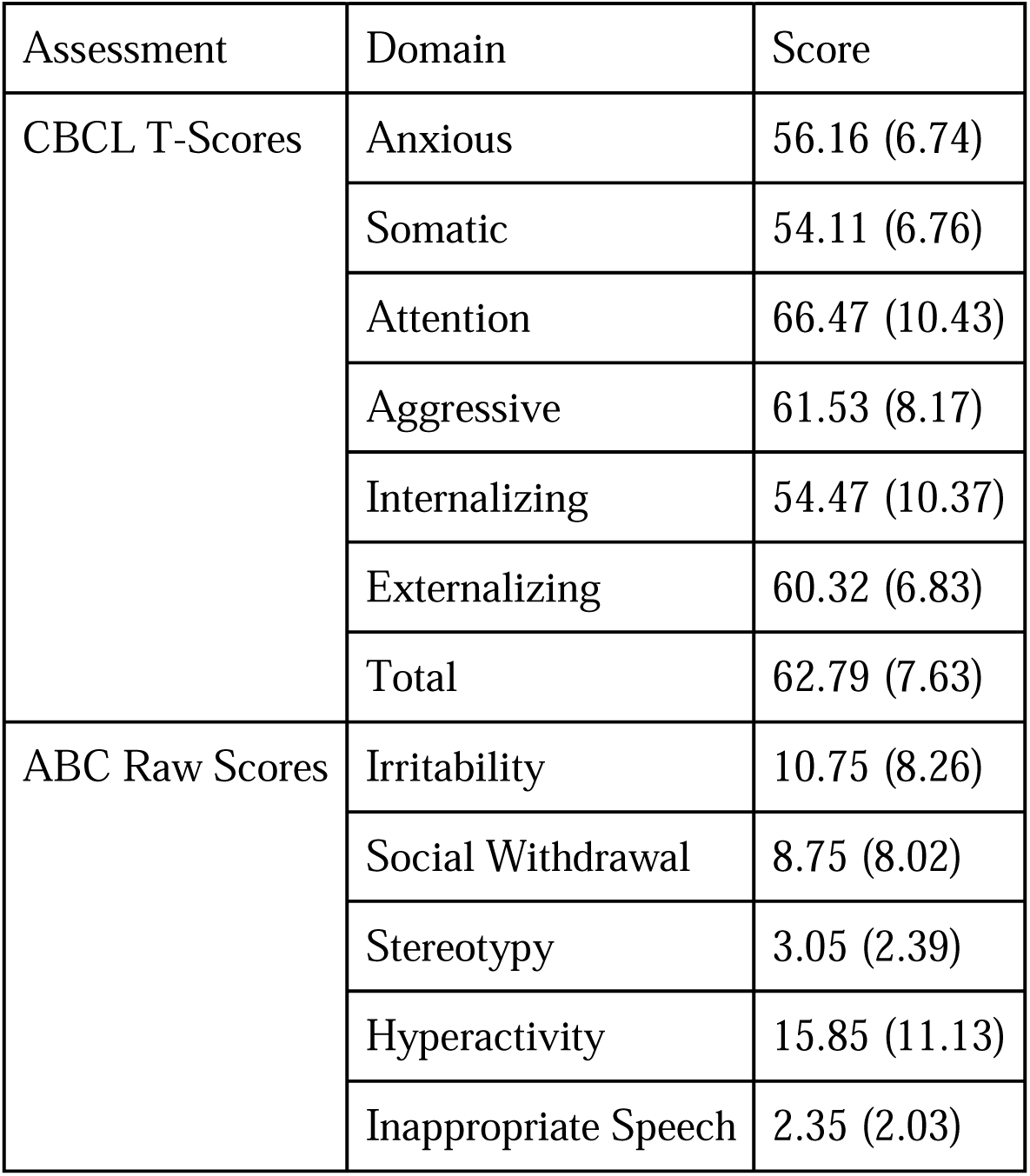
Behavioral Survey Results. CBCL scores are in T Scores with a population mean of 50 and standard deviation of 10; ABC scores are raw scores. Maximum scores for each domain are 45, 48, 21, 48, and 12, respectively, for Irritability, Social Withdrawal, Stereotypy, Hyperactivity, and Inappropriate Speech.

#### Externalizing Symptoms

A history of aggression was reported in 15/20 participants. In four cases aggressive behavior resolved over time. In six cases, aggression improved but remained present at some level, and in two cases aggression remained consistent. In four cases, aggression worsened over time, particularly as children became physically stronger. Daily violent outbursts were reported in one case. A history of irritability was reported in 16/20 participants. Two parents reported irritability resolved over time, five reported improvement, and four reported consistent symptoms over time. Three parents reported irritability increased with age and these were the same individuals who experienced increased aggression.

#### ADHD Symptoms

A history of inattention was reported in 18/20 participants and reportedly improved over time in 15 participants. Inattentive symptoms lessened in 12 of those cases. Four parents reported consistent levels of inattention over time. Hyperactivity was reported in 17/20 with four parents reporting hyperactivity was no longer present post-puberty, 10 reporting hyperactivity improved but was still present, and three reported consistent symptoms over time. Overall, attention problems were reported as less prominent when individuals aged out of the school system and demands to attend for extended periods of time or in specific conditions (i.e., desk work) decreased. Thirteen carried a formal ADHD diagnosis, and most received medication (see below).

#### Autism Traits

Eight participants carried a previous diagnosis of autism spectrum disorder. Socially, almost all parents described their children as motivated and interested in interacting with close family and familiar people outside of the family. Most reported at least one reciprocal friendship (range 1-15; median and mode = 5), however, caregivers routinely mentioned that their children struggled to independently maintain friendships without adult support due to communication and/or behavioral problems. Regarding nonverbal communication, some parents reported intact skills (i.e., appropriate eye contact, gesture, facial expression) while others reported inconsistent nonverbal skills. Most participants gestured spontaneously, responded to others’ facial expressions and tone of voice, and effectively used eye contact when motivated to communicate. A few parents reported a lack of gestures and flat expressions or expressions not always appropriate to the context. Regarding communication, pragmatics (i.e., the social use of language) were frequently impaired. Communication was largely related to expressing wants and needs and sharing information about personal interests. Difficulty staying on topic, sustaining back and forth conversation, or conversing about topics outside of one’s interests were frequently reported. Importantly, improvement in expressive language and pragmatics was reported even post-puberty, albeit still delayed relative to developmental expectations. There were reports of decreased social motivation following puberty in two participants.

Repetitive and restricted behaviors (RRBs) and interests were prominent, even in those who did not carry an autism diagnosis. All parents reported a history of RRBs, which largely persisted over time. One parent reported symptoms improved although remained to a lesser degree, one reported RRBs worsened over time and the rest remained consistent. Insistence on sameness and inflexible adherence to routines and rituals were common. Examples included collecting objects (e.g., phone cases, cars, sneakers, baby dolls), eating the same meal for years, insisting on wearing shirts with pockets, changing clothes multiple times throughout the day, extreme layering of clothing and accessories, watching the same part of a show or movie over and over again, carrying around particular objects, compulsively shutting open doors or insistence on turning books pages one by one, lining up cars, sorting cards, and verbal rituals including those requiring family members to respond in a particular manner. There were some reports of stereotyped speech, although few reports of motor stereotypies. Sensory symptoms were also common. The SAND-Interview indicated high levels of sensory reactivity differences characterized by seeking (7.0 ± 3.6), hyperreactivity (5.7 ± 3.5), and hyporeactivity (5.0 ± 3.3). A high threshold for pain and temperature was the most endorsed item (17/20), followed by seeking pressure/mouthing objects (14/20), being startled or bothered by sounds (12/20), fascination with certain sounds (e.g., specific parts of a song or show; 12/20), and resisting touch (10/20). Nail picking and biting was reported in most patients as well as picking other parts of the skin or scabs. In some cases, nail picking was very severe (e.g., picking fingernails until they bleed).

#### Other Psychiatric Symptoms

Mania and psychotic episodes were reported in one participant who carried a diagnosis of bipolar disorder (diagnosed with a mood disorder and later bipolar disorder during adolescence), and in one participant whose parents suspected bipolar disorder starting in early adolescence. Similarly, both participants had a history of aggression, one of which subsided with medication. The parents of both individuals described the presence of “imaginary friends;” in one case emerging during early childhood with more added over time and in the other first emerging during the teenage years. One patient was verbally fluent and the other spoke in phrases. Parents both described fluctuating periods of growth and plateaus. Both were female and were described as socially motivated with areas of adaptive strength (e.g., toilet trained, able to write).

#### Findings from Standardized Caregiver-Report Questionnaires

This section examines results from caregiver-reported questionnaires compared to parent-reported symptoms gathered from the semi-structured clinical interview with a licensed clinician (Table 3). The CBCL/ABCL did not pick up elevated or clinically significant internalizing symptoms in this cohort but did pick up on externalizing problems including aggressive behavior and attention problems/ADHD. At the item level, the ABC adequately captured distractibility (19/20), impulsivity (18/20), and inattentive (17/20) symptoms. Finally, on the Short Sensory Profile, parents reported above average sensory avoidance (11/19) and sensory sensitivity (13/19).

### Medication History

Medications for ADHD symptoms were used by 11 of the 13 individuals with an ADHD diagnosis. The average age participants began taking these medications was 8.45 ± 2.8 years (range 4-12). Participants tried an average of three ADHD medications prior to current medications or stopping ADHD medication altogether. Nine participants still took ADHD medication at the time of evaluation. Methylphenidate (e.g., Ritalin, Concerta, Quillivant) was the most common ADHD medication, which was prescribed for nine participants, however five stopped due to adverse reactions (e.g., increased skin picking, irritability, rebound impulsivity). Guanfacine was prescribed for five participants, all five of whom reported ongoing treatment. Mixed amphetamine salts (Adderall) was used for three participants and stopped for two because of adverse reactions (e.g., increased problem behaviors, skin picking, irritability). Three participants were actively taking clonidine for ADHD with no adverse effects.

Ten participants had a history of medication use for anxiety, mood, or behavioral problems, with nine under active treatment. The average age participants began taking these medications was 12.8 ± 3.8 years (range 5-18). The most common medications was sertraline (Zoloft) which was prescribed in five participants, three with ongoing therapy and two stopped due to adverse effects (e.g., aggressive behavior, crying, agitation). Quetiapine (Seroquel) was tried for three participants and stopped in two cases due to ineffectiveness. An additional three patients used fluoxetine (Prozac); none discontinued this medicine. Aripiprazole (Abilify) was prescribed to one participant and discontinued due to weight gain.

Seven participants took medication for sleep, six with ongoing treatment. No medications were stopped due to adverse effects, although Clomipramine (Anafranil) and eszopiclone (Lunesta) were stopped by one participant due to ineffectiveness. Ongoing medications for sleep included guanfacine (n=2), clonidine (n=3), and melatonin (n=2).

Three participants had a history of anti-seizure medication (ASM) use and two were on ASMs for seizure prevention at the time of evaluation. No seizure medications were stopped due to adverse effects.

Three participants used oral contraceptives for menstrual management. Three participants took medication for gastrointestinal issues including reflux (lansoprazole), gastritis (omeprazole), and constipation (polyethylene glycol). Two participants took medication for seasonal allergies, one of which also took medication for asthma. Other medical concerns requiring medication included dermatitis (n=1), type 2 diabetes (n=1), arthritis (n=1), and enlarged prostate (n=1).

**Table 3a.**
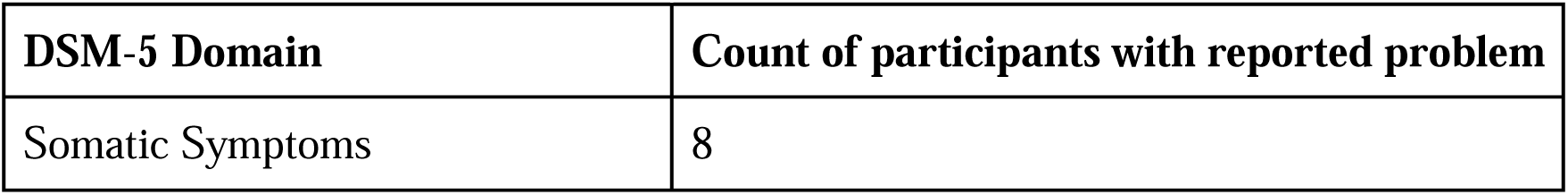
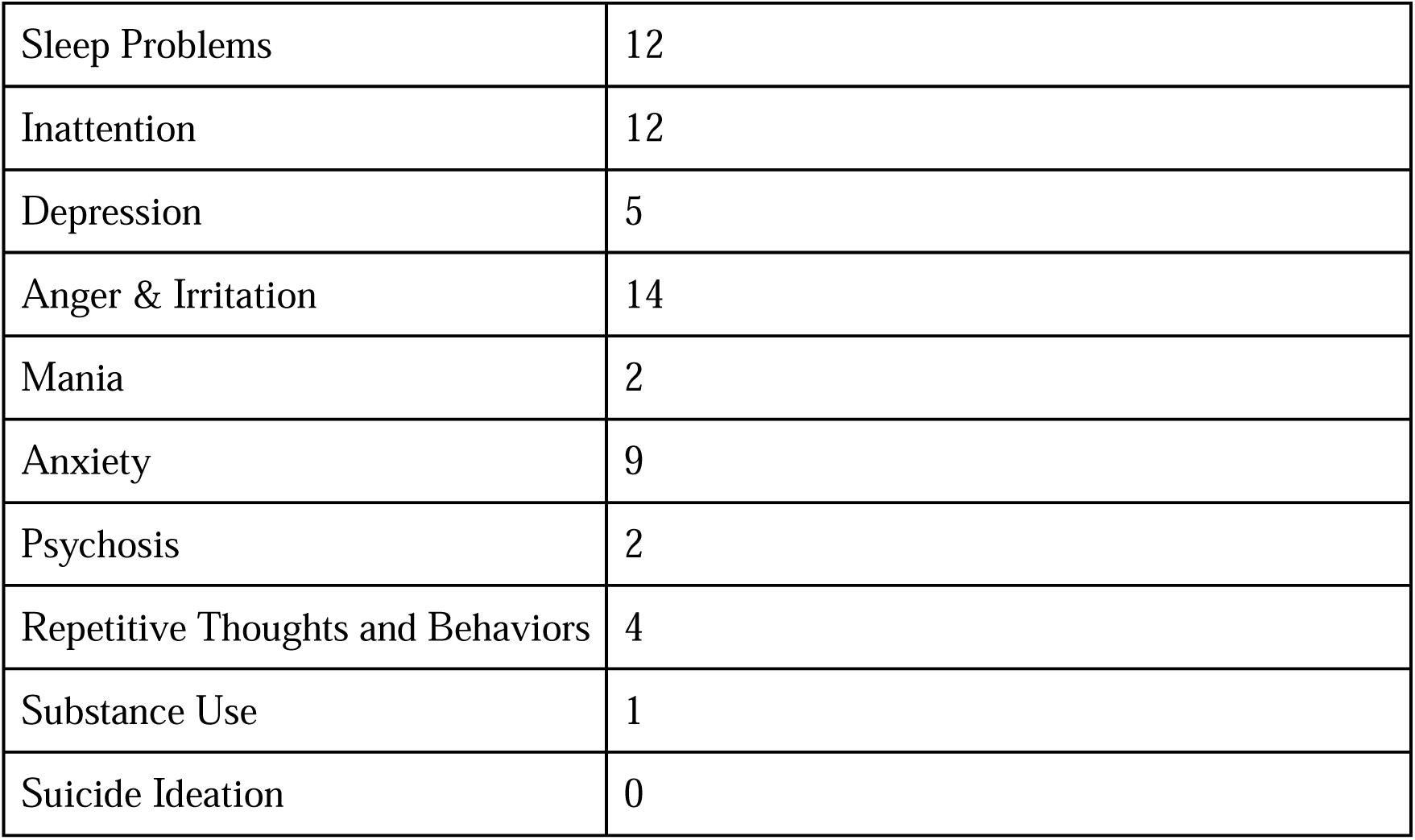
DSM-5 Cross Cutting Interview Results. Count of caregivers who reported behaviors in each domain as a ‘1: Slight, Rare, less than a day or two’ or higher. Caregivers reported based on the previous two weeks of time.

### Visual Analog Scale: Top Concerns

Caregivers reported their top three concerns for their children during childhood and rated them on a scale from 0 (least concerned) to 100 (most concerned) based on (1) level of concern during childhood and (2) current level of concern (Figure 3). Concerns were categorized into nine groups: ADHD features (e.g., ‘focus’, ‘hyperactivity’), Anxiety, Cognition (e.g., “intelligence”, “cognitive awareness”), General Development (e.g., “global delays”, “reaching milestones”), Independence (e.g., “ability to learn life skills”, “ability to live independently”), Medical/Health (e.g., “low muscle tone”, “seizures”), Problem Behavior (e.g., “aggression”, “biting others”), Social (e.g., “play with peers”, “having friends”), and Speech/Communication (e.g., "language development”, “speech”).

**Figure 3.**
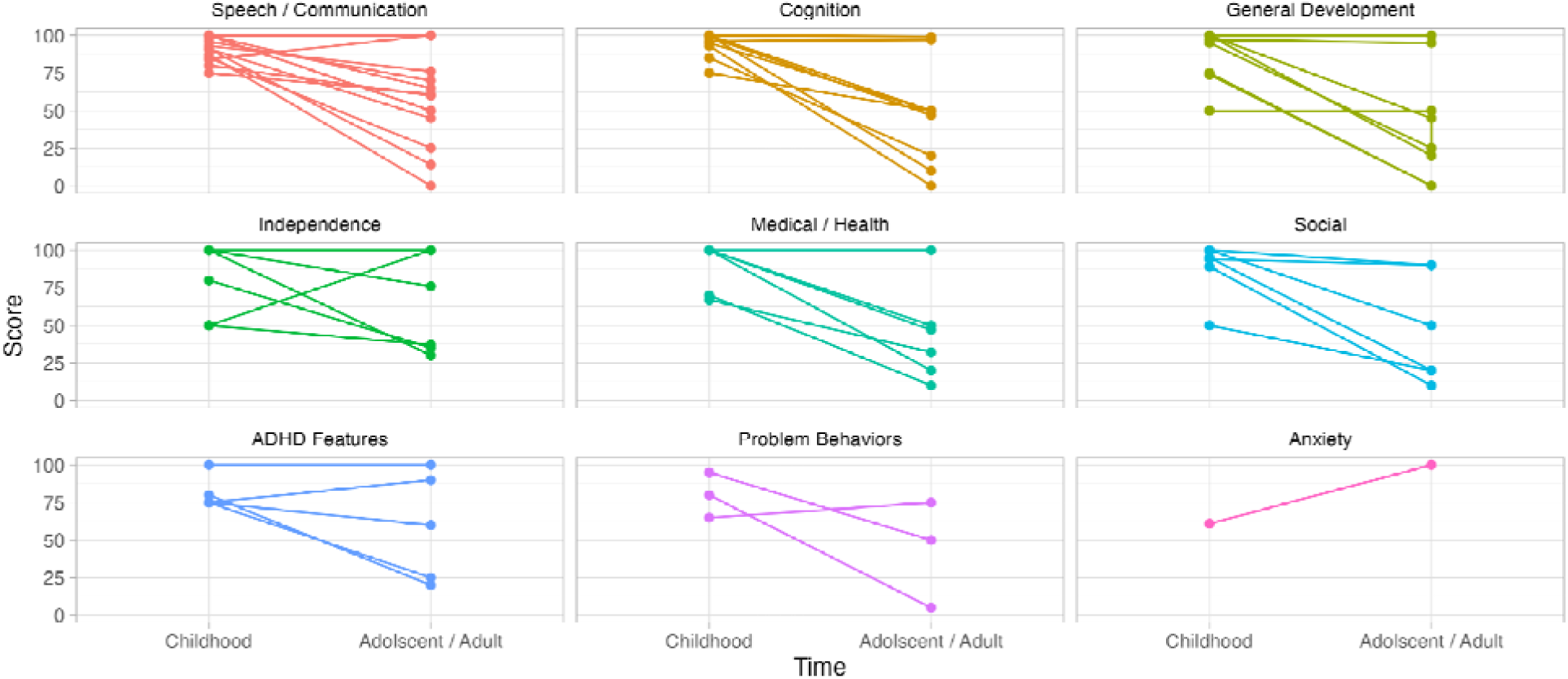
Caregiver top childhood concerns. Caregivers listed their top three concerns for their children in childhood; they rated them on a scale of 0-100 twice, once for how concerned they were in childhood and once for how concerned they were currently, in adolescence/adulthood. Concerns were grouped into categories. Categories are shown in order of most commonly to least commonly reported.

Speech and communication challenges were the most common childhood concern (15/20), with an average rating of 92 ± 8, this concern fell to an average score of 58 ± 30 in adulthood.

Nine caregivers endorsed concerns related to cognition in childhood with an average rating of 94 ± 9 which fell to 47 ± 35 in adulthood. Nine expressed concerns related to general development in childhood with an average rating of 82 ± 21, which fell to 40 ± 37 in adulthood. Caregivers of six participants reported concerns relating to independence with an average rating of 80 ± 25 during childhood and 63 ± 33 in adulthood. Six caregivers reported concerns relating to medical or health related issues with an average rating of 90 ± 15 in childhood, which fell to an average rating of 43 ± 32 in adulthood. Social concerns were reported by six caregivers, with an average rating of 88 ± 19 in childhood and 47 ± 36 in adulthood. Five concerns related to ADHD had an average score of 81 ± 11 in childhood and 59 ± 36 in adulthood. Three caregivers reported problem behaviors as a top concern in childhood with an average rating of 80 ± 15, which fell to 43 ± 35 in adulthood. Lastly, one caregiver reported anxiety as a concern in childhood with a score of 61 which increased to 100 in adulthood.

Caregivers then reported the top three concerns for their children as adults and rated them on a scale from 0 (least concerned) to 100 (most concerned) (Figure 4). Concerns were grouped into 12 categories. The most common concern was related to independence (e.g., “taking care of herself”, “daily living tasks”) and had an average score of 86 ± 15. This was followed by concerns related to socialization which was rated at 76 ± 17. Next, problem behaviors were reported by 8 caregivers (“hitting”, “verbal and physical aggression”) and were rated at 78 ± 8. Five rated speech/communication as a top concern, with an average rating of 91 ± 10. Four caregivers reported concerns related to housing (e.g., “long term care,” “future housing”) with an average rating of 96 ± 7. Three reported concerns related to participant’s health with an average rating of 78 ± 20, and two related to the caregiver’s health with an average rating of 98 ± 3. Two top concerns fell in each of these categories: ADHD features, anxiety, cognition, and financial security with average ratings of 99 ± 1, 83 ± 14, 82 ± 17, and 83 ± 11, respectively. Lastly, one concern related to general development was reported at 80.

**Figure 4.**
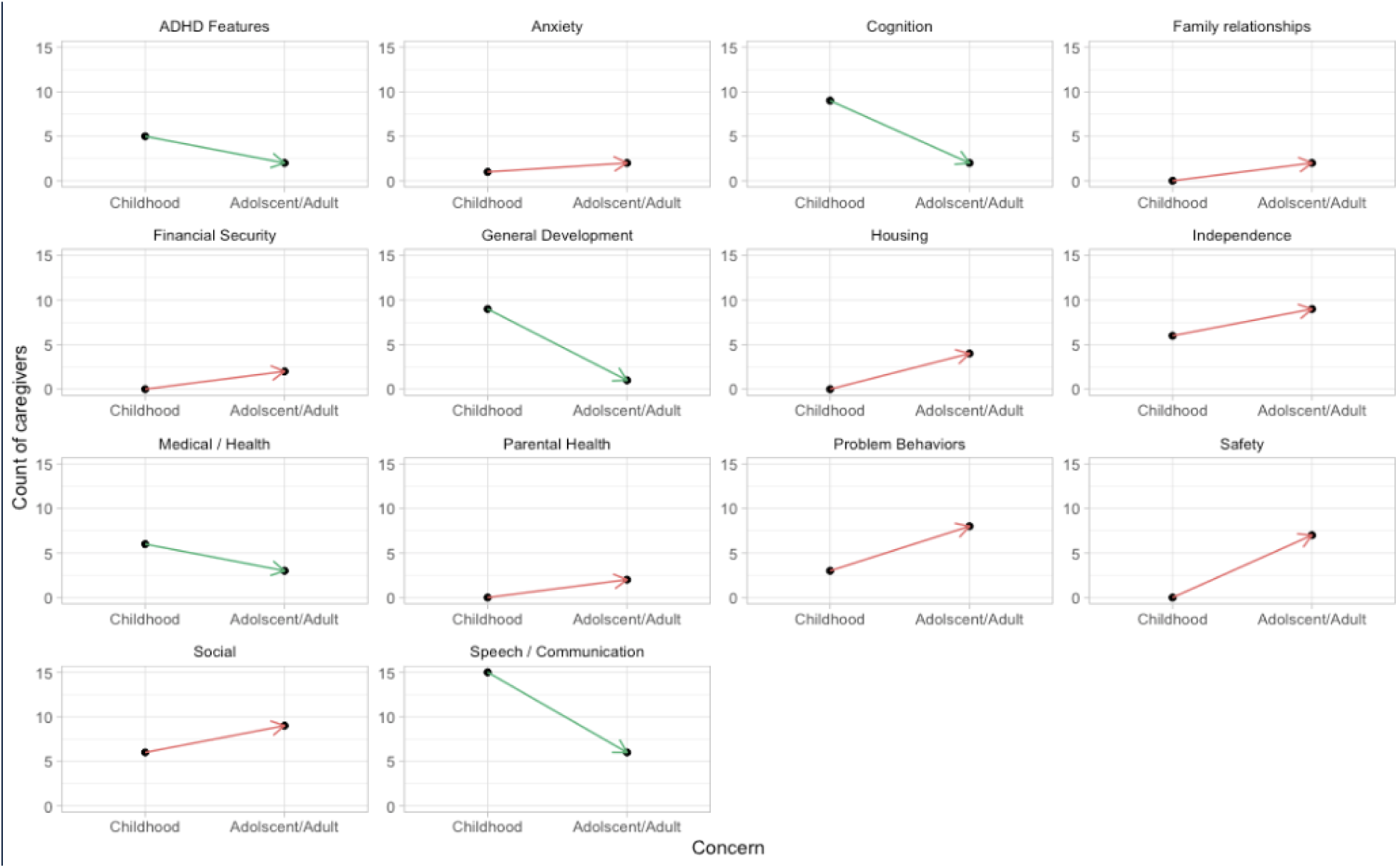
Change in top caregiver concerns. Caregivers were asked the top three concerns they had for their children in childhood and the top three concerns they had for their children in adolescence/adulthood. The concerns were grouped into categories. The count of concerns within each category was summed in childhood and adolescence/adulthood. Green arrows indicate if fewer caregivers noted top concerns in this domain across time, while red arrows indicate that more caregivers reported a concern in this domain in adulthood as compared to childhood. Categories are in alphabetical order.

## DISCUSSION

Prior literature describing FOXP1 syndrome focuses primarily on childhood symptomatology, including medical and neurobehavioral features of the disorder. Previous prospective phenotyping at our center revealed high rates of ADHD, anxiety, and autism traits including repetitive behaviors and sensory symptoms (10, 11). Other publications identified key medical concerns such as sleep and ocular problems (15). Psychiatric manifestations in adolescents and adults with FOXP1 syndrome had yet to be examined and is of critical interest given psychiatric decompensation and regression reported in other genetic neurodevelopmental disorders. To further understand the post-pubertal FOXP1 syndrome phenotype, data was obtained from parents or caregivers of 20 individuals with FOXP1 syndrome who had already started puberty based on Tanner scales. Characterization included medical, developmental, adaptive, and behavioral measures. Detailed clinical interviews assessed psychiatric and medical symptoms and how those developed or changed over time. Lastly, we compared information obtained from semi-structured clinical interviews to standardized scores from validated questionnaires to assess survey effectiveness in this cohort.

Early developmental delays were reported across participants. All walked independently by 2.5 years old. All spoke using at least single words, with 19 speaking in phrases and 11 speaking in sentences. Fluent speech developed later than expected, on average, by age 7 and parents reported continued improvement in the complexity of language even following puberty. Articulation problems and difficulty with intelligibility were commonly reported as were pragmatic difficulties. No skill loss was reported.

Regarding adaptive functioning, toilet training was a challenge for all but one participant. While most participants had generally achieved continence, accidents were reported even in young adults, one who carried a change of clothes to their job. Vineland-3 Socialization domain scores were relatively stronger than Communication and Daily Living Skills scores. In the Communication domain, v-sores were relatively stronger for receptive language compared to expressive language, although mean age equivalents were similar and at the 3-4 year old level. Interestingly, written language skills were, on average, at almost the 7-year-old level with many participants demonstrating some capacity to read and write. Personal care skills were also identified as an area of weakness. While many individuals participated in activities of daily living (e.g., dressing, showering, teeth brushing, food prep), there was heavy reliance on prompting or reminders.

Regarding educational and vocational history, about half of participants were enrolled in a mainstream class for preschool and by the end of elementary school all had transitioned to special education classes or required modified curriculums/support. Home schooling was used intermittently by some families who could not find proper educational placements. In terms of vocational achievement, five had paying jobs, and most caregivers of younger participants reported a desire for their child to hold a job in the future. None of those with paying jobs were able to support themselves and obtaining jobs required support from caregivers. Most unemployed individuals who had aged out of the school system required substantial support.

In terms of psychiatric history, anxiety was commonly reported (17/20) and over half carried a formal anxiety disorder diagnosis. In contrast, depression was only reported in one participant. Externalizing features were highly prevalent with a history of aggression and/or irritability reported in most participants. Aggressive features tended to lessen over time, with symptoms in 10/15 participants having resolved or lessened with age. In cases where aggression persisted over time, behavior was often severe and frequent. ADHD symptoms of inattention and hyperactivity were reported in most participants. In the majority of cases, hyperactivity improved, and in some cases resolved, with age. This is consistent with findings from ADHD studies in the general population (30). Psychopharmacological treatment was used by at least half the cohort to target symptoms of anxiety, mood, behavior, and ADHD. Participants had mixed effects of medications, and many required multiple trials due to varying side effects. Fluoxetine (i.e., Prozac) was the only medication for mood features not discontinued due to side effects (n=3). Guanfacine (n=5) and Clonidine (n=3) were successfully used for ADHD symptoms without adverse effects. While this cohort is too small to form recommendations about medication use, preliminary data suggest some medications may be better tolerated than others, and multiple medication trials in this population is typical.

Psychotic symptoms were reported in two female participants with first episodes during the early teenage years. One carried a bipolar disorder diagnosis. The prevalence of symptoms of bipolar disorder (suspected or diagnosed) in this group was therefore 10% compared to the national prevalence of ∼3-4% (31, 32). This cohort was underpowered to assess if true prevalence is increased in FOXP1 syndrome and these findings warrant continued investigation. Importantly, the two individuals with symptoms of bipolar disorder also had better adaptive and language skills compared to the group. There was no history of catatonia or other forms of psychiatric decompensation.

In terms of autism spectrum disorder, forty percent carried the diagnosis, however, several parents reported their child’s presentation was atypical due to social strengths. Social motivation and relationships with peers and familiar family members or adults were frequently described. Caregivers reported weaknesses in pragmatic communication, such as reciprocal conversation. Despite social interest, many participants required adult support to maintain social relationships and to plan social activities with peers. Restricted and repetitive behaviors were present across participants and a persistent feature. Behaviors such as collecting objects, rituals, and compulsive behaviors were reported to impact functioning and engagement in activities of daily living. Sensory symptoms were endorsed by most caregivers as well, including, sensory seeking, hyperreactivity (i.e., noise and touch sensitivity), and hyporeactivity (i.e., high pain threshold). Nail picking and biting was reported in most participants and in certain cases was very severe. A high pain threshold was the most commonly reported item on the SAND, which is consistent with reports from other genetic syndromes associated with autism (33–35)

Data obtained from structured and semi-structured clinical interviews were compared to standardized parent report questionnaires. The Vineland-3 maladaptive behavior internalizing and externalizing domains accurately picked up on elevated symptoms, although lack diagnostic specificity. Conversely, the CBCL/ABCL only captured externalizing behaviors and did not reveal elevated levels of internalizing features, likely due to poor applicability of questions for individuals with language deficits. ABC T-score cutoffs were poor indicators of symptomatology and raw scores were more useful. These findings are worthy of consideration when determining clinical outcome assessments for future studies and, ultimately for clinical trial design.

Finally, results from the VAS revealed the most common concerns in childhood were speech/language and cognition and the degree of concern for both dropped considerably with age. In adolescence/adulthood, the most common concerns shifted to independence and socialization. As parents accepted communication and cognitive challenges they were perceived as less concerning. Problem behaviors were more often reported as a concern for parents in adolescence/adulthood than in childhood, indicating that even with reported reduction in many problem behaviors with age, those that remain, significantly impair quality of life. Our findings corroborate a previous study which identified cognition, communication, and behavior problems as top concerns for parents (15).

## Conclusions

Taken together, results are reassuring, with many families reporting their children continued to gain skills, particularly related to increased independence in communication and personal care well past puberty. There were no reports of developmental regression, neuropsychiatric decompensation or catatonia. Psychotic symptoms in two patients suggest there may be an increase relative to the general population and warrants continued study. Although larger studies are needed, relative to other rare genetic neurodevelopmental syndromes, FOXP1 syndrome does not appear to be associated with devastating regression and psychiatric changes emerging during and after puberty. The most commonly reported challenges were anxiety, externalizing behaviors (ADHD, irritability, aggression), repetitive behaviors, and difficulties with continence (i.e., continued accidents even when largely independent at toileting). Aggressive behavior requires additional study given the severity of symptoms in certain individuals creating safety concerns within the home. Optimistically, the most reported symptoms in this post-pubertal FOXP1 syndrome cohort can all be targeted using existing interventions, and only about half of participants were actively being treated. Findings from this study support the need for natural history studies to elucidate developmental trajectories prospectively and to further assess optimal endpoints for use in future clinical trials.

## List of abbreviations

ABC: Aberrant Behavior Checklist
ABCL: Adult Behavior Checklist
ADHD: Attention-deficit/hyperactivity disorder
ASD: Autism spectrum disorder
ASM: Anti-seizure medication
CBCL: Child Behavior Checklist
CIPIPID: Caregiver Interview for Psychiatric Illness in Persons with ID
DSM-5: Diagnostic and Statistical Manual of Mental Disorders Fifth Edition
Early Skills: Early Skill Attainment and Loss
FOXP1: Forkhead box protein P1
ID: Intellectual disability
PMS: Phelan-McDermid syndrome
RRB: Repetitive and restricted behaviors
SAND: Sensory Assessment for Neurodevelopmental Disorders
SP-2: Short Sensory Profile 2
VAS: Visual Analog Scale
Vineland-3: Vineland Adaptive Behavior Scales, 3rd Edition - Comprehensive Interview Form

## Additional Files

Additional File 1. Visual Analog Scale

## DECLARATIONS

### Ethics approval and consent to participate

This study was approved by Mount Sinai’s Program for the Protection of Human Subjects. Informed consent was obtained from parents or legal guardians.

## Consent for publication

n/a

## Availability of data and materials

The datasets used and/or analyzed during the current study are available from the corresponding author on reasonable request.

## Competing interests

TL receives funding from the Phelan-McDermid Syndrome Foundation and is on the advisory board of the CHAMP1 research foundation.

AK is on the Advisory Board for the Klingenstein Third Generation Foundation, Ovid Therapeutics, David Lynch Foundation, ADNP Kids Research Foundation, and Ritrova Therapeutics and consults to Acadia, Alkermes, Jaguar Therapeutics, GW Pharmaceuticals, Neuren Pharmaceuticals, Clinilabs Drug Development Corporation, Scioto Biosciences, and Biogen.

PMS is a developer of the Sensory Assessment for Neurodevelopmental Disorders, which is licensed by Mount Sinai to Stoelting, Co.

JDB consults to BridgeBio, holds a patent for IGF-1 in Phelan-McDermid syndrome, holds an honorary professorship from Aarhus University Denmark, receives research support from Takeda and Oryzon, and is a journal editor for Springer Nature.

## Author Contributions

TL was involved in the conceptualization, design, data collection, analysis, and preparation of the manuscript. HS was involved in the data collection and analysis. RS was involved in data collection. AR was involved in data collection. AK was involved in design and preparation of the manuscript. JDB was involved in conceptualization and preparation of the manuscript. PMS was involved in conceptualization, design, data collection, analysis, and preparation of the manuscript.

## Data Availability

All data produced in the present study are available upon reasonable request to the authors.

## Acknowledgements

We would like to thank all of the wonderful individuals who participated in this study for their time and dedication to improve our understanding of FOXP1 syndrome and to the Beatrice and Samuel A. Seaver Foundation for funding this study.

